# Modelling NHS 111 demand for primary care services: a discrete event simulation

**DOI:** 10.1101/2023.05.22.23290330

**Authors:** Richard Pilbery, Madeleine Smith, Jonathan Green, Daniel Chalk, Colin O’Keeffe

## Abstract

**Background:** Almost half of the 16,650,745 calls to NHS 111 each year are triaged to a primary care disposition. However, there is evidence that contact with a primary care service occurs in less than 50% of cases and triage time frames are frequently not met. This can result in increased utilisation of other healthcare services.

This feasibility study aimed to model *in-silico* the current healthcare system for patients triaged to a primary care disposition and determine the effect of reconfiguring the system to ensure a timely primary care service contact.

**Methods:** Data from the Connected Yorkshire research database were used to develop a model and Discrete Event Simulation in Python, using the SimPy package. This included all 111 calls made in 2021 by callers registered with a Bradford GP who were triaged to a primary care disposition, and their subsequent healthcare system access during the following 72 hours.

We simulated 100 runs of one year of 111 calls and calculated the mean difference and 95% confidence intervals in primary care contacts, emergency ambulance (999) calls and avoidable ED attendances.

**Results:** The simulation of the current system estimated that there would be 39,283 (95%CI 39,237–39,328) primary care contacts, 2,042 (95%CI 2,032–2,051) 999 calls and 1,120 (95%CI 1,114–1,127) avoidable ED attendances. Modifying the model to ensure a timely primary care response resulted in a mean increase in primary care contacts of 37,748 (95%CI 37,667–37,829), a mean reduction in 999 calls of -449 (95%CI -461– -436) and a mean reduction in avoidable ED attendance of -26 (95%CI -35– -17).

**Conclusion:** In this simulated study, ensuring timely contact with a primary care service would lead to a significant reduction in 999 and 111 calls, and ED attendances (although not avoidable ED attendance). However, this is likely to be impractical given the need to almost double current primary care service provision. Further economic and qualitative research is needed to determine whether this intervention would be cost effective and acceptable to both patients and primary care clinicians.

## Introduction

The National Health Service (NHS) 111 service aims to assist members of the public with urgent medical care needs and is the successor to the NHS Direct service in England. Following pilots in four sites it was rolled out nationally, with the final site going live in England in 2014, and in 2019/20 received over 19 million calls (NHS England, 2020). Its key founding objective was to provide easy access to support for the public with urgent care needs, to ensure they received the “right care, from the right person, in the right place, at the right time” (UK Government, 2011). It is also the key component of the 24/7 Integrated Urgent Care Service outlined in the NHS Long Term Plan (England, 2019).

Approximately 55% of all calls made to the English National Health Service (NHS) 111 call service are triaged to a referral to a primary care service. If a timely service cannot be provided to patients, it is possible that this will result in patients calling 999 or attending emergency departments (ED) directly. We have previously reported that just under half (47.6%) of callers to 111 triaged to a primary care service disposition made contact with a primary care service as their first healthcare interaction (Pilbery et al., 2023). There was evidence that patients more frequently attended an ED when they had not had previous contact with a primary care service. However, rates of avoidable attendance were similar whether callers had contact with a primary care service prior to attendance at ED or not.

Other studies examining caller behaviour and the impacts on the wider healthcare system (Lewis et al., 2021; Nakubulwa et al., 2022), have utilised datasets from 2017 and earlier, and patterns of activity identified then, may not be applicable now.

This study aimed to develop a Discrete Event Simulation to model the current healthcare system for callers triaged to a primary care disposition. Discrete Event Simulation models allow for the simulation of queuing problems, in which entities (such as patients) queue for processes that require resources (Caro et al., 2016). Our primary objective was to develop a model which represented the current healthcare system in relation to callers to 111 who received a primary care service disposition following triage. Our secondary objective was to use the model to estimate the predicted impact and associated resourcing required to ensure a timely primary care contact for all callers to the 111 service in a region of Yorkshire.

## Methods

### Conceptual modelling

Prior to developing a simulation that represents the trajectories of patients within the model, it is necessary to develop a conceptual model: an appropriately simplified collection of assumptions about the components and structure of the ‘real-world’ (Banks et al., 2014) that can act as a blue print for the in-silico model development. This technique has its origins in soft systems methodology (Checkland, 1999), and enabled us to work with stakeholders in the ambulance service, NHS 111, general practice and emergency departments to co-produce processes and capture their knowledge of the health and social care system (Figure 1). This co-production is not only crucial in terms of improving the accuracy of the model, but also an important component of stakeholder engagement and the formation of strategic partnerships (Green, 2018).

**Figure 1:**
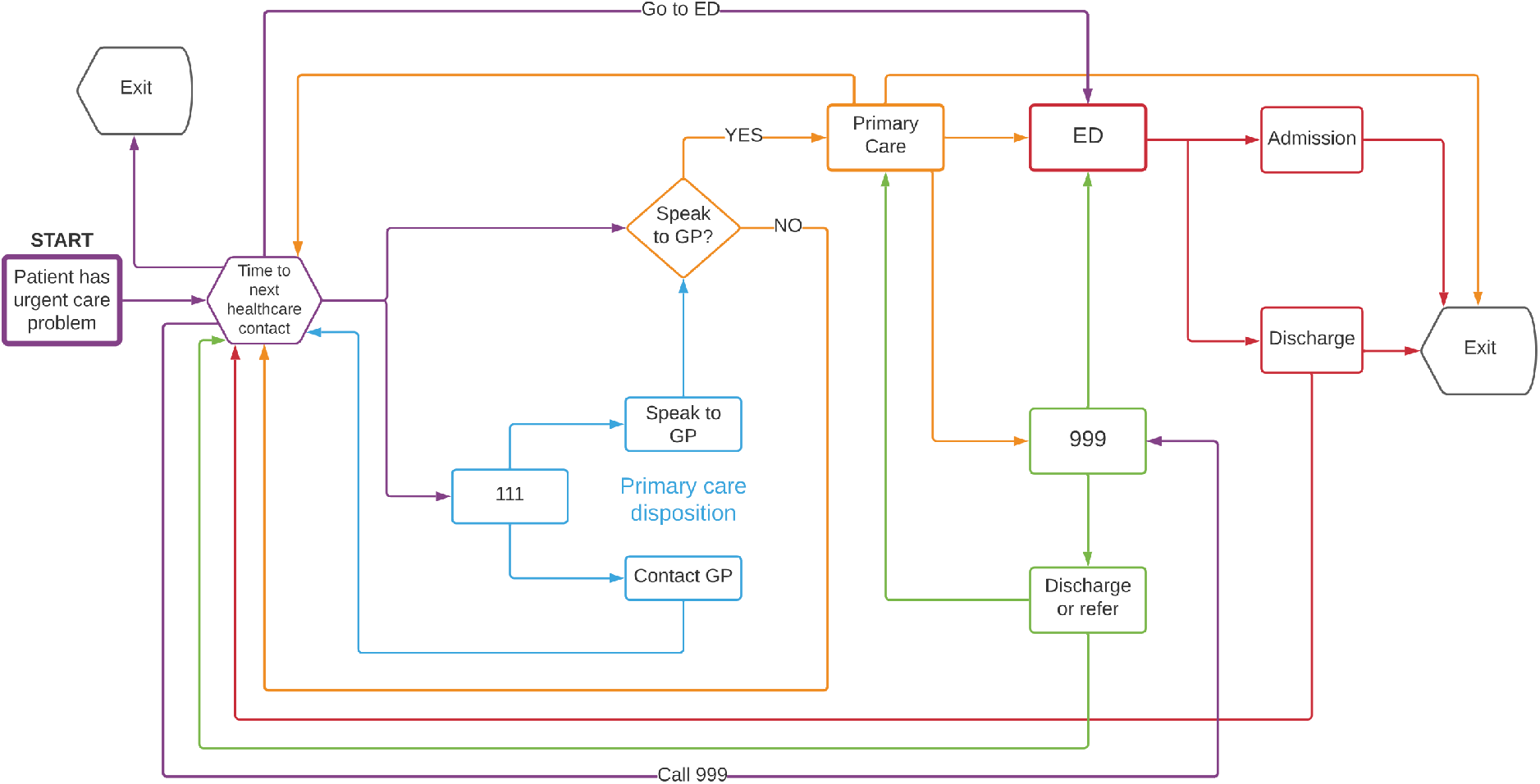
Design-orientated conceptual model of patients with an urgent care problem

### Data

In tandem with the conceptual modelling phase, we obtained routine, retrospective data from the Connected Yorkshire research database, which provides linked data for approximately 1.2 million citizens across the Bradford and Airedale region of Yorkshire. Datasets include 111 and emergency ambulance (999) call data, as well as primary and secondary care (including emergency department and in-patient activity).

We identified all 111 calls between the 1st January 2021 and 31st December 2021 for patients who were triaged to a primary care disposition and registered with a General Practitioner (GP) in the Bradford area at the time of the call. Subsequent healthcare system access in the following 72 hours following the first (index) call was identified by searching datasets containing details of 111 and 999 calls, and primary care, hospital emergency department and in-patient admissions (Pilbery et al., 2023).

### Discrete event simulation

Using Brennan et al. (2006) taxonomy, we determined that Discrete Event Simulation (DES) was a good choice of operational research methodology to address the research question. DES is a flexible modelling technique that can represent complex behaviour of callers to 111, and their interactions with the healthcare system simultaneously. DES can also incorporate capacity and resource constraints and account for chance, for example the stochastic behaviour of callers (Caro et al., 2016; Karnon et al., 2012; Pitt et al., 2016). DES is also useful, because it makes it possible to ask ‘what if’ questions such as the research question. It has been used widely in healthcare, examining health care systems operations, and disease progression, screening and health behaviour modelling (Forbus & Berleant, 2022; Vázquez-Serrano et al., 2021).

We created an *in-silico* version of the conceptual model using the programming language Python (v3.9.6) and the SymPy library (v4.0.1, Meurer et al., 2017). In addition, we created a web-based dashboard in order to present the results of the simulation and provide an opportunity for users/stakeholders to run their own simulations. To achieve this, we used Plotly Dash (v5.11.0, Plotly Technologies Inc, 2015) and created a fully contained environment in docker (v4.2.0.7078) which is available from the study GitHub repository (https://github.com/RichardPilbery/MOOOD-study).

Patients enter the model at the point they are triaged to a primary care disposition by 111 and have exited the service. The ‘arrival’ rate of callers into the model is determined by sampling from an exponential probability distribution with lambda set to the 1/mean arrival rate for 111 calls, stratified by hour, yearly quarter and whether the call occurred at a weekend (Supplementary 1). When a patient is created in the simulation, they are assigned a primary care disposition with an associated triage acuity (Figure 2). This consists of a time frame within which a contact with a primary care service should be made, and specifies whether the contact should be face-to-face or remotely, for example by telephone. The allocation is random, but weighted according to the proportions seen in the Connected Yorkshire data. Once the patient has been allocated a primary care disposition, the uniform distribution is sampled to determine whether the primary care contact will be achieved in the specified triage time frame. This is assumed to be the case when the sampled value is less than proportion calculated from the Connected Yorkshire dataset.

**Figure 2:**
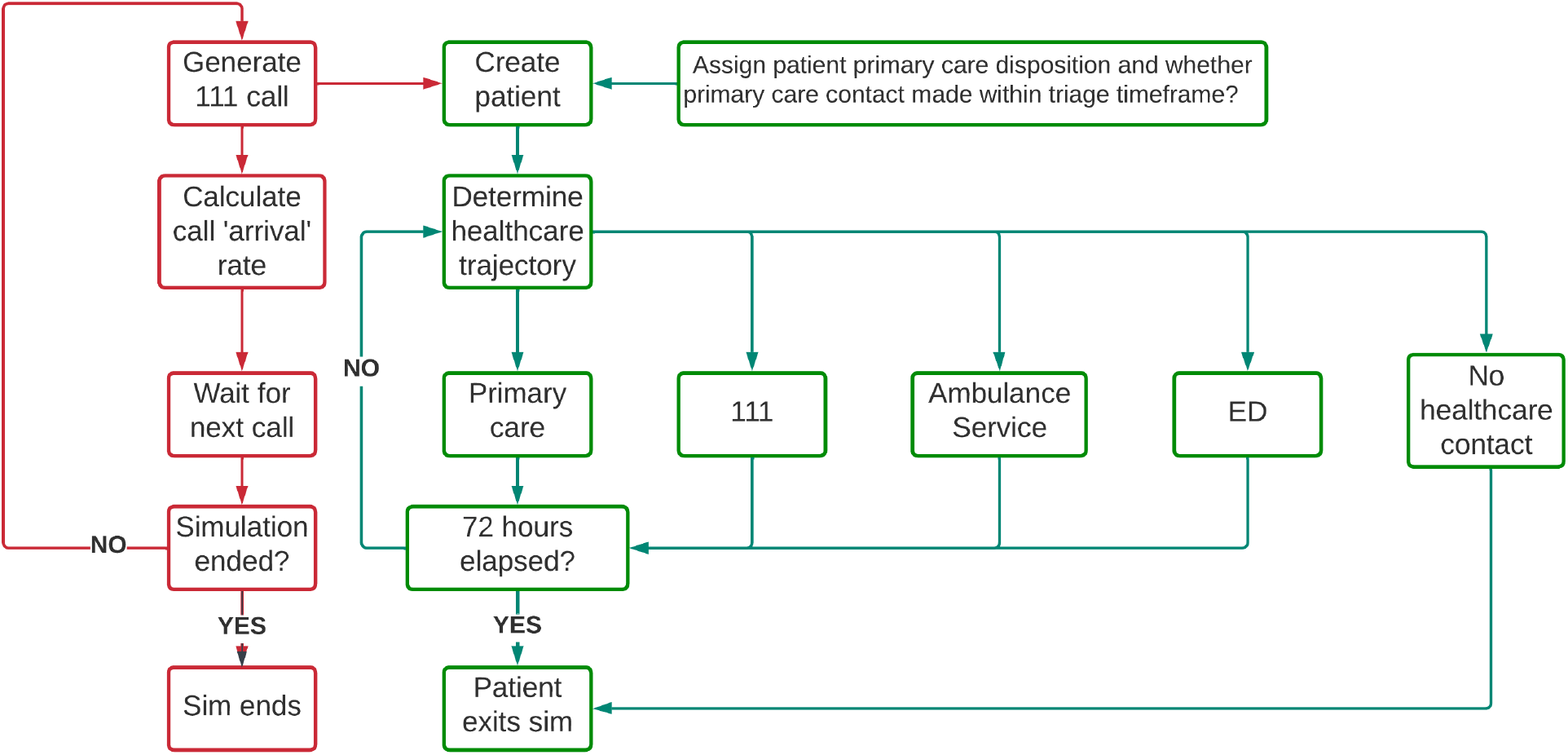
Programmatic simulation flow chart

Patient trajectory through the healthcare system is determined using transition probabilities calculated from the Connected Yorkshire dataset using the statistics software, R (R Core Team, 2022). The next health care service (including no further contact) is determined by random sampling, weighted by whether a primary care service contact was made immediately after the index 111 call, and the yearly quarter (Supplementary 2). If the next healthcare service access is determined to be the emergency department (ED), then the uniform distribution is sampled to determine whether the admission will be avoidable. As before, this is assumed to be the case when the sampled value is less than proportion calculated from the Connected Yorkshire dataset. The proportions are stratified by primary care disposition and whether initial contact was made with a primary care service.

For this study, an ED attendance is classed as being ‘avoidable’ when it meets the O’Keeffe et al. (2018) criteria. They defined an avoidable attendance as a patient presenting to a consultant-led ED which provides a 24 hour service with full resuscitation facilities and designated accommodation for the reception of emergency care patients (referred to as a type 1 ED (NHS Digital, 2023)), but who do not receive investigations, treatments or referral that required the facilities of a type 1 ED.

Unless the patient is allocated to ‘no further healthcare contact’, in which case the patient exits the simulation, the ‘queue’ time i.e. the time the patient waits until the service is accessed and the time that the healthcare activity takes, is calculated. The ‘queue’ time is calculated by sampling from a distribution specific to the current healthcare service being access and the next, for example index 111 call to emergency department attendance (Supplementary 3). Activity times are calculated by sampling from distributions specific to the service being accessed (Supplementary 4). In both cases, the optimal distribution was chosen by the Python library fitter (v1.5.2, Cokelaer et al., 2023) based on the distribution with the lowest sum of square errors.

Patients remain in the simulation until either they are allocated to ‘no further healthcare contact’ or the elapsed time exceeds 72 hours. The model runs for a period of 1 year of simulated time.

### Analysis

Once the model was developed, we performed 100 runs of 1 year’s worth of simulated 111 calls and monitored the patient’s trajectory over the subsequent 72 hours. As the model is stochastic, multiple runs of the model were necessary to ensure that results are representative and not due to stochastic noise. A summary of all simulations was reported as the mean and 95% confidence intervals for counts and proportions.

Since all models are approximations of the ‘real world’, we used the historic data provided by the Connected Yorkshire dataset to see how closely our model matches with real data (the primary objective), visually and descriptively analysing the difference in temporal distribution of calls between the actual and simulated data, as well as comparing quarterly aggregated healthcare service access by patients, following the index 111 call. Finally, we performed a visual assessment of patient trajectory, to determine whether it approximated actual patient behaviour.

For the secondary objective, we undertook the ‘what if’ analysis, examining the hypothetical situation whereby all index 111 calls with a primary care disposition received a timely (i.e. within the specified call triage time) response from a primary care service. No capacity constraints were placed on primary care services in order to estimate the true resourcing that would be required to meet the demand appropriately. We calculated the mean difference and 95% confidence intervals in healthcare system usage between simulations and determined the difference in mean proportion of avoidable admissions for callers who presented to an ED.

### Ethical approval

This study was approved by the Bradford Learning Health System Board in accordance with the Connected Yorkshire NHS Research Ethics Committee (REC) and Confidentiality Advisory Group (CAG) approvals relating to the Connected Yorkshire research database (17/EM/0254). No separate Health Research Authority (HRA) approval was required for this study.

### PPI

The application and protocol for this study was review by the Yorkshire Ambulance Service NHS Trust patient research ambassador. In addition, Connected Bradford have an active patient and public involvement group and a representative attended the approvals board for this study.

## Results

111 call activity in both simulations was similar to the actual call activity seen in the real data, although overall, the simulation did slightly overestimate the number of calls, particularly between 00:00 and 06:00(Figure 3).

**Figure 3:**
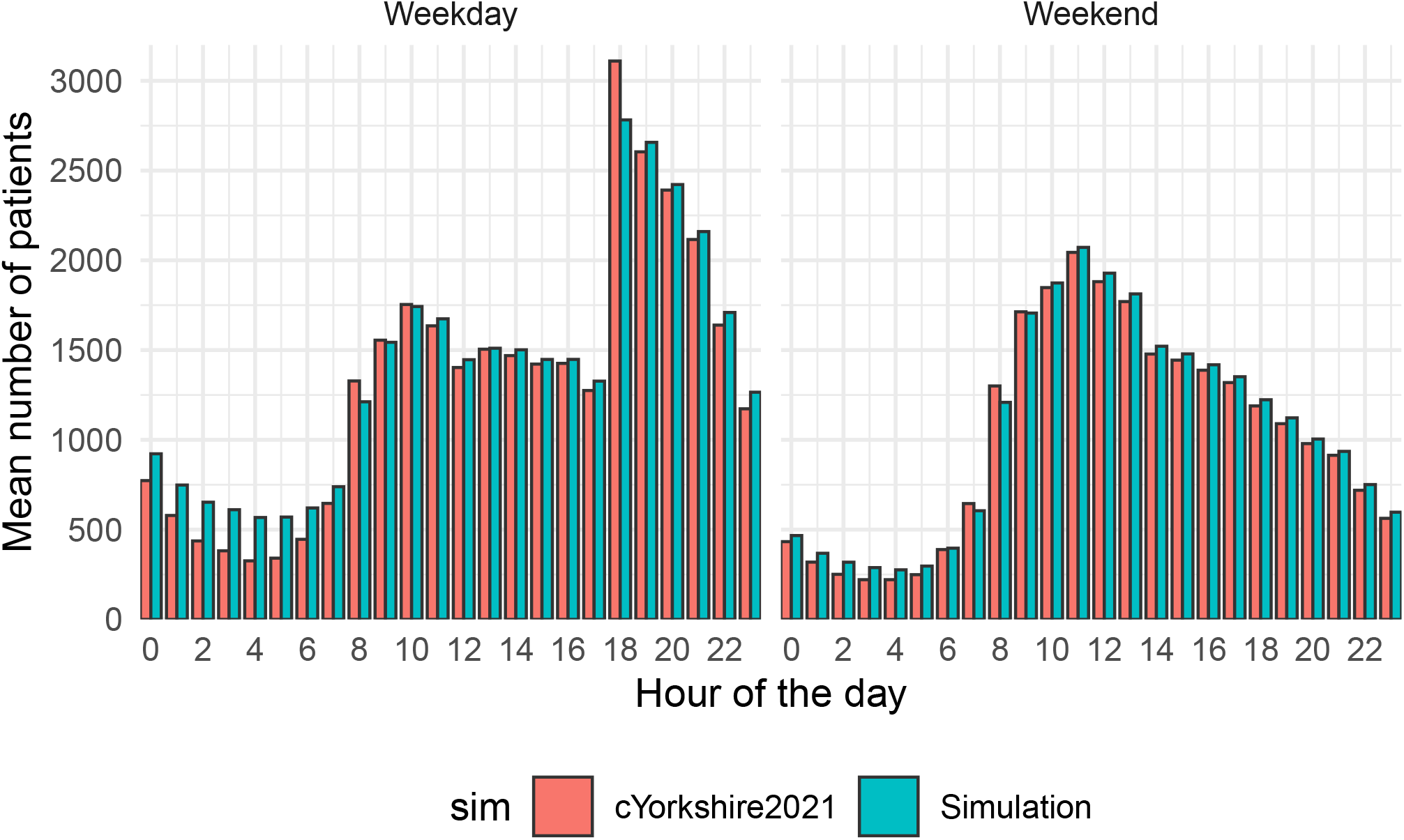
111 call volume by hour and day of week

### Healthcare system access

Overall, the simulation tended to underestimate 999 call activity and overestimate all other healthcare service activity. Primary care, emergency department admission and subsequent 111 call activity was most closely simulated (Table 1, Figure 4).

**Table 1:** Comparison of simulation activity with Connected Yorkshire data

| Healthcare service | cYorkshire2021 (N) | Simulation (N, 95%CI) |
| --- | --- | --- |
| Ambulance service | 2,623 | 2,042 ( 2,032– 2,051) |
| Emergency department | 9,290 | 9,337 ( 9,319– 9,355) |
| Primary care | 38,680 | 39,283 (39,237–39,328) |
| In-patient | 4,367 | 4,668 ( 4,654– 4,681) |
| 111 | 4,710 | 4,789 ( 4,771– 4,806) |

**Figure 4:**
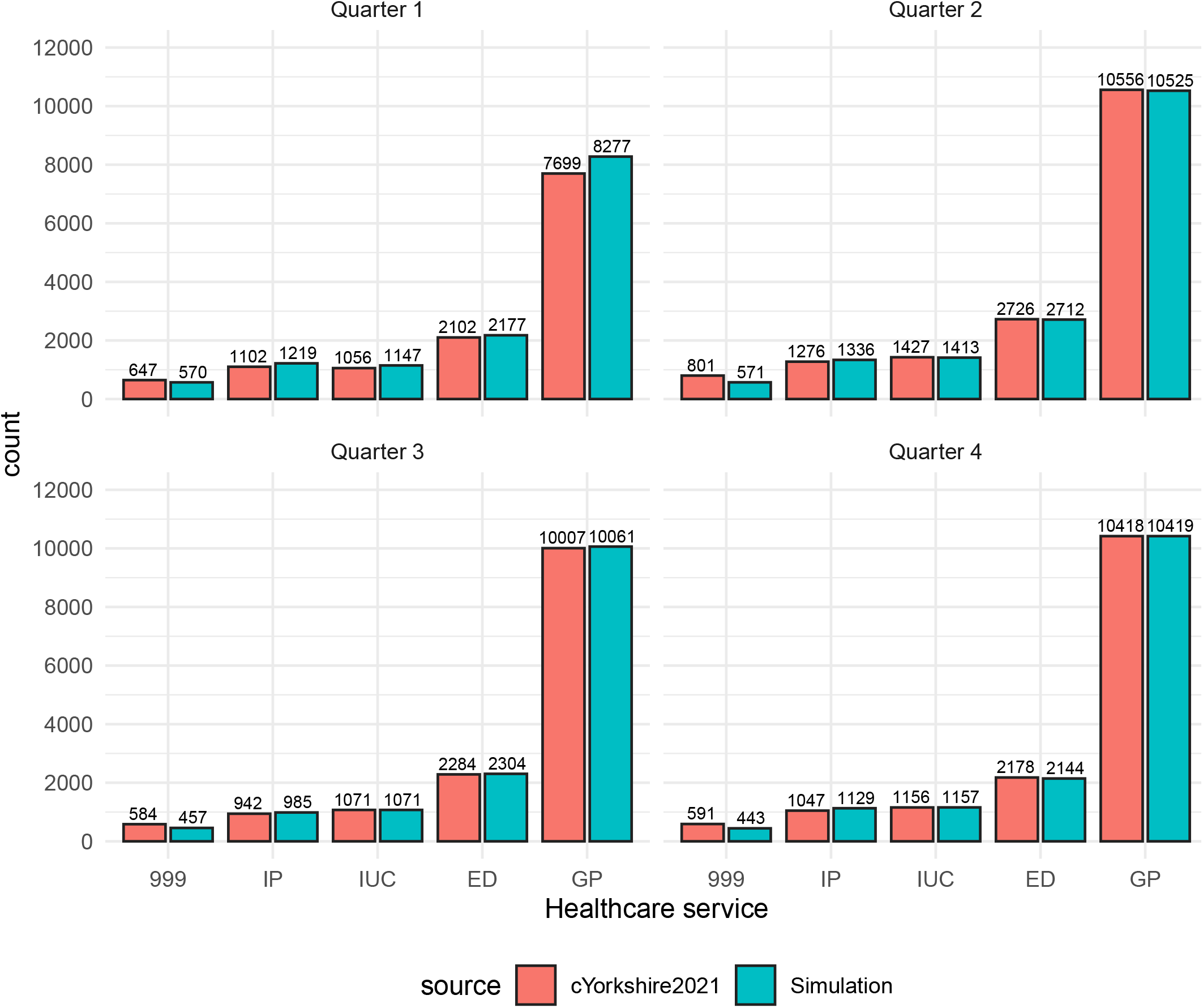
Healthcare system activity, stratified by data type and yearly quarter

### “What if?” scenario

Assuming no constraints on primary care service ability, we simulated a hypothetical scenario whereby all 111 calls with a primary care disposition received a timely primary care contact (Table 2).

**Table 2:** Comparison between simulation and what if scenario

| Service | Simulation (mean, 95%CI) | What if (mean, 95%CI) | Mean difference (mean 95%CI) | Mean proportion change (mean %, 95%CI) |
| --- | --- | --- | --- | --- |
| Ambulance service | 2,042 ( 2,032–2,051) | 1,593 ( 1,584–1,602) | -449 ( -461– -436) | -22 (-30.2– -13.8) |
| Emergency department | 9,337 ( 9,319–9,355) | 8,228 ( 8,213–8,244) | -1,109 (-1,133– -1,085) | -11.9 (-18.3– -5.5) |
| Primary care | 39,283 (39,237–39,328) | 77,030 (76,964–77,097) | 37,748 (37,667–37,829) | 96.1 (92.2– 99.9) |
| In-patient | 4,668 ( 4,654–4,681) | 4,620 ( 4,607–4,632) | -48 ( -66– -30) | -1 (-3– 1) |
| 111 | 4,789 ( 4,771–4,806) | 3,074 ( 3,061–3,087) | -1,715 (-1,737– -1,693) | -35.8 (-45.3– -26.3) |

### ED attendance

In the original Connected Yorkshire dataset, there were 9,290 emergency department attendances and 1,029 (11.1%) met the O’Keeffe et al. (2018) definition of an avoidable attendance. The simulated dataset estimated a mean ED attendance of 9,337 cases (95%CI 9,319–9,356), of which, 1,120 (95%CI 1,114–1,127; mean proportion, 12%, 95%CI 5.6–18.4%) were classed as avoidable. In the ‘what if’ scenario, there was a reduction of more than 10% in ED attendances, but little change in the proportion of avoidable attendance (Table 3).

**Table 3:**
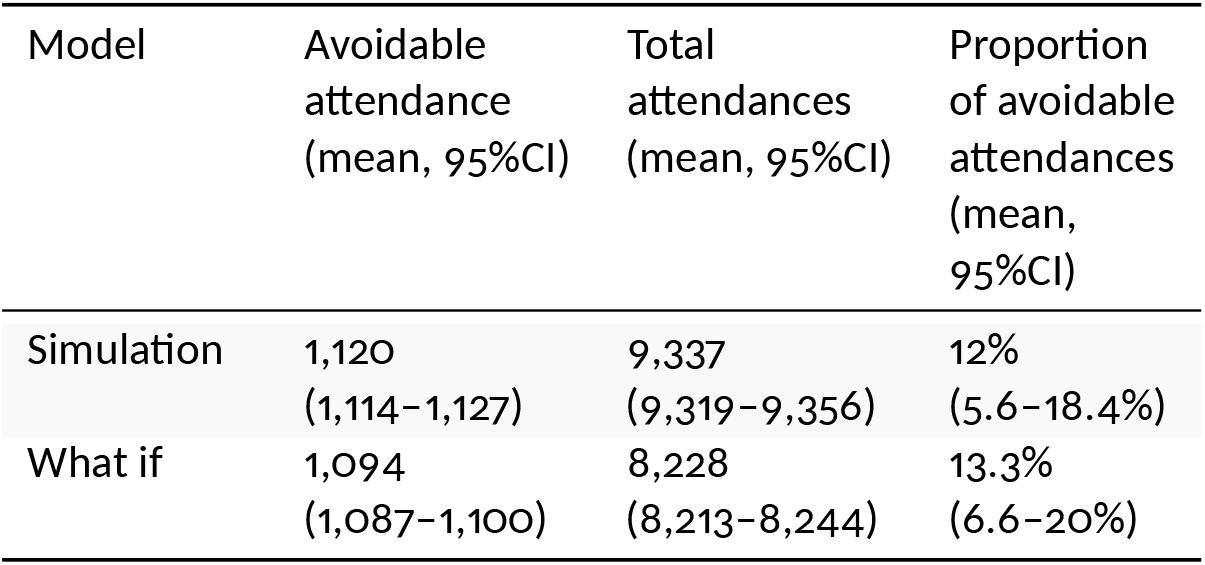
Simulated ED attendance rates stratified by avoidable admission

| Model | Avoidable attendance (mean, 95%CI) | Total attendances (mean, 95%CI) | Proportion of avoidable attendances (mean, 95%CI) |
| --- | --- | --- | --- |
| Simulation | 1,120 (1,114–1,127) | 9,337 (9,319–9,356) | 12% (5.6–18.4%) |
| What if | 1,094 (1,087–1,100) | 8,228 (8,213–8,244) | 13.3% (6.6–20%) |

## Discussion

Using a simple simulation model, we have been able to successfully capture the healthcare system as it relates to callers to 111 who are triaged to a primary care disposition. We used the model to estimate the impact of a hypothetical scenario in which primary care services had sufficient capacity to provide a service to all 111 callers triaged to their service and that all callers elected to make contact with the service. We determined that the ‘base case’ model sufficiently closely replicated the real world data to be confident that its estimates of the impact of the scenario tested would be useful.

In the simulated ‘what if?’ scenario, we found that ensuring a timely primary care contact for all 111 callers triaged to a primary care disposition, would lead to a significant reduction in ambulance service and 111 contacts and emergency department admissions. However, to achieve this it would be necessary to almost double the number of primary care contacts provided by the current healthcare system which, in the wider context of clinical staff shortages in general practice (British Medical Association, 2023; The Health Foundation, 2022) and the NHS (Waitzman, 2022), is unlikely to be feasible. While we did not conduct an economic analysis, the data suggests that the savings made by reducing certain healthcare service contacts are unlikely to offset the additional cost in providing additional primary care services.

Even if the capacity issues could be overcome, adjusting caller behaviour could be even more challenging. Almost 39% of callers in the base case model did not seek any further healthcare contact in the 72 hours after the index 111 call. Previous qualitative work on understanding why patients call an ambulance for ‘primary care problems’ highlighted the perceived limitations of out-of-hours primary care services by patients to address their needs and, in the case of patients with chronic conditions, previous negative experiences of community-based healthcare services being unable to ‘help’ would need to be addressed (Booker et al., 2014). In addition, there are reasons why patients may not follow advice to contact primary care which are beyond the control of the health system, such as work or family/childcare commitments, transport issues (including weather-related) and demographic factors such as younger age, female sex and low socio-economic background (Parsons et al., 2021); factors that would be unaffected by changes in service provision. That said, there is limited evidence that increasing availability of primary care services (such as offering extended hours) can lead to a reduction in access other services. This is particularly pertinent, since most 111 calls with a primary care disposition occur out-of-hours (Whittaker et al., 2016).

This work has generated stakeholder discussion about whether the apparent supply/demand problem identified by the simulation could be addressed by utilising the Additional Roles Reimbursement Scheme. In addition, they have also suggested that further qualitative work might help to understand which primary care roles best suit particular caller presentations (for example a consultation with physiotherapist rather than a GP for a musculoskeletal presentation), which may reduce the proportion of callers who do not access healthcare services following the 111 call and are potentially left with an unmet healthcare need.

### Strengths and weaknesses

This study benefits from utilisation of a robust method (discrete event simulation) that has been widely used and accepted as a method to simulate healthcare system changes (Caro et al., 2016). In addition, the model was informed by relevant stakeholders and a linked dataset. However, it is nevertheless a simple working model, and does not take account of factors that have previously been associated with inappropriate ED attendance, such as age, sex, ethnicity and clinical input into the 111 call (Nakubulwa et al., 2022).

The model may benefit from more extensive validation and verification than the simple ‘black box’ validation described here, in which outputs from the model were compared with the equivalent real world metrics. Additional validation measures, such as performing sensitivity analyses to determine the impact of inaccuracies in modelling assumptions could be useful in increasing confidence in the model’s predictions (Macdonald & Strachan, 2001). In addition, verification measures such as extensive review of code by others can help to identify issues in the software engineering side of the development (Raunak & Olsen, 2014). However, the model and its source code has been made available free and open source for others to interrogate and develop, and the simple validation steps we have taken give us relative confidence in our estimates of the scale of the resourcing required to meet demand.

The data that informed the model only covers a discrete region in West Yorkshire, which may affect generalisability. Bradford is mainly an urban area and the 13th most deprived local authority in England (out of 333) based on the Index of Multiple Deprivation (City of Bradford Metropolitan District Council, 2019). In addition, it is based on data collected in 2021, during the pandemic, which may not be representative of the current healthcare trajectory for patients who call 111. However, the open source nature of the model means that others can use the model to parameterise according to the data from their own regions.

Further economic and qualitative analysis would be beneficial to determine whether an intervention such as this would be cost-effective, feasible and acceptable to patients.

## Conclusion

In this simulated study, ensuring timely contact with a primary care service is estimated to lead to a significant reduction in 999 and 111 calls, and ED attendances (although not avoidable ED attendance). However, this is likely to be impractical given the estimated need to almost double current primary care service provision. Further economic and qualitative research is needed to determine whether this intervention would be cost effective and acceptable to both patients and primary care providers.

## Data Availability

The complete model, including transition probabilities and inter-arrival, activity and queue distributions are available from the study GitHub repository (https://github.com/RichardPilbery/MOOOD-study). The software is supplied under a GNU General Public License.

## Acknowledgements

This work uses data provided by patients and collected by the NHS as part of their care and support. The authors would also like to thank Dr. Eithne Cummins and Dr. Jon Dickson for providing a stakeholder perspective to the study, and Dr. Hazel Squires for early support in operational research methodology. In addition, we are grateful for the support provided by the team at Connected Yorkshire, especially Kuldeep Sohal and John Birkinshaw.

The work was supported as a project of the NIHR Applied Research Collaboration for the South West Peninsula Health Service Modelling Associates (HSMA) Programme (https://sites.google.com/nihr.ac.uk/hsma) which also provided the training required in Python and Discrete Event Simulation.

## Contributors

RP and DC conceived and designed the study. RP obtained the research approvals to access the datasets and acts as guarantor for the paper. RP and MS developed the code necessary to extract the data that informed the model. RP coded the model. DC provided mentoring support and guidance for the project, and delivered the training in Python and SimPy. CO assisted with the design of the study. All authors drafted the manuscript and contributed substantially to its revision.

## Funding

This paper presents independent research by the NIHR Applied Research Collaboration Yorkshire and Humber (ARC YH). This work was supported by the National Institute for Health Research Applied Research Collaboration South West Peninsula and Yorkshire and Humber.

## Disclaimer

The views expressed in this publication are those of the author(s) and not necessarily those of the National Institute for Health Research or the Department of Health and Social Care.

## Notes

### Competing Interest Statement

The authors have declared no competing interest.

### Author Declarations

This study was approved by the Bradford Learning Health System Board in accordance with the Connected Yorkshire NHS Research Ethics Committee (REC) and Confidentiality Advisory Group (CAG) approvals relating to the Connected Yorkshire research database (17/EM/0254).

